# Health indicators as measures of individual health status, perceived importance, and their associated factors—an observational study

**DOI:** 10.1101/2022.08.28.22279311

**Authors:** Xia Jing, Yuchun Zhou, Temiloluwa Sokoya, Sebastian Diaz, Timothy Law, Lina Himawan, Francisca Lekey, Lu Shi, Sarah Griffin, Ronald W. Gimbel

**Author notes:** Corresponding Author Xia Jing, MD, PhD, Edwards Hall 519, Department of Public Health Sciences, College of Behavioral, Social, and Health Sciences, Clemson University, Clemson, SC 29634.

## Abstract

**Background:** Self-rated health status, a subjective measure, is used broadly to describe an individual’s overall health status. Our long-term goal is to create a more objective, comprehensive, and accurate measure of individual health status. We selected 29 health indicators and prioritized them by conducting online surveys. Thirteen of these 29 indicators received relatively more consistent ratings across 3 samples.

**Objectives:** To explore the main and interaction effects of 4 demographic factors as independent variables (age, gender, professional group, and educational level) in the importance ratings of the 13 health indicators.

**Methods:** We conducted a 4-way multivariate analysis of variance (MANOVA) with *post-hoc* testing to examine the effects of independent variables on all 13 dependent variables. Descriptive statistics and bivariate correlation analysis were also conducted.

**Design:** Cross-sectional study.

**Setting:** An online survey (≥ 18 years).

**Participants:** 791 participants in the USA.

**Results:** 13 health indicators were significantly correlated with each other. Age correlated with most of the health indicators (8 of 13). The MANOVA modeling results indicated that gender, age, and education levels significantly affected the combination of the 13 health indicators. There was a significant interaction effect by age and professional group on 5 health indicators.

**Conclusions:** Age is critical in rating the 13 health indicators. Among all the statistically significant main effects of demographic factors, the effect sizes descend regarding age, gender, educational level, and interaction between age and professional group. These results can provide a foundation for further studies to explore behavioral interventions for individual subgroups.

**Article Summary:** 

**Strengths and limitations of this study:**

- The work establishes the interactions and effects between demographic data (age, gender, education, and professional group) and the perceived importance of 13 health indicators via MANOVA analysis
- The interactions and effects of demographic data on the importance ratings of the 13 health indicators
- can guide future study designs for behavioral interventions
- Deep analysis of the demographic variables and their effects on and interactions with the rating results are helpful for thoroughly understanding the perspectives
- The study is an observational study despite with relatively large sample size and a robust analysis
- The data are not racially representative

## Introduction

Health indicators can be a useful tool in various healthcare scenarios and public health. Usage examples include measuring individual health status [1], measuring populational health status [2], tracking the outcomes of preventive services over time, and comparing health statuses between focused populations or among different locations [2–4]. Disease stages have long been used to help physicians determine more appropriate treatment plans and to form relatively more homogeneous patient groups to make them comparable within groups. The most commonly used measurement of overall individual health status is self-rated health status, which has been shown to have good validity among the United States population [3,5,6], although the measure was initially used to predict mortality among the elderly. Self-rated health status has also been broadly used by the World Health Organization and the Organisation for Economic Co-operation and Development. Nevertheless, there are no clear criteria for each level of self-rated health status; thus, the rating might be subjective due to the lack of clear criteria. This subjectivity has been recognized as a potential limitation, which could affect the comparison among different population groups [3].

Due to the lack of existing measures, more objective, comprehensive, and accurate health indicators to measure individual health status are needed, although life expectancy, morbidity, and mortality have all long been used to measure group health outcomes. We conducted a literature review [1,3,7–11] to compile 29 health indicators [12] that can measure individual health status. We then conducted a pilot study among 4 commercial electronic health record (EHR) systems to determine how many of such health indicators were included in each system. The results showed that no system includes *<u>all</u>* health indicators [13]. From a practical point of view, it can be burdensome for EHR users to collect all 29 health indicators [14]. We, therefore, conducted a public perspective survey to ask the general public to rate the importance of each health indicator, aiming to use the rating results to further prioritize health indicators. The survey results showed that, across 3 samples, the respondents agreed most on 13 health indicators among the 29 [12]. These 13 health indicators were immunization, personal care needs, cancer screening, HIV testing, self-rated health status, air quality index >100, dentist supply, health literacy, blood sugar level, blood triglycerides, high-density (HDL) and low-density lipoprotein (LDL), and total cholesterol.

In this paper, we further analyzed the responses of the public perspectives for any relationships between the rating results of these 13 health indicators and their demographic information to better understand the public rating results for these health indicators by age, gender, education, and professional group. The detailed analysis of these variables could aid future study designs for health-related behavioral interventions in various subgroups of the general public.

## Methods

The study was a Cross-sectional study. We used an online survey among the general public who are older than 18 years old to obtain their importance ratings on the 29 health indicators derived from the literature review.

This study is a follow-up project on the exploration of public perspectives on 29 individual health indicators. Our previous study indicated that 13 of the 29 health indicators received more consistent ratings across 3 samples: Ohio University, ResearchMatch [15], and Clemson University [12]. In this study, we examined these 13 health indicators according to demographic factor, including age, gender, educational level, and professional group. The analysis did not include race or ethnicity, mainly due to imbalanced data [12].

We recoded the demographic information as follows. Age included 3 groups: group 1 (age ≤ 35 years), group 2 (age 36–55 years), and group 3 (age ≥ 56 years). Gender included group 1 (females) and group 2 (males). Eighteen respondents selected “transgender” or “prefer not to answer.” Due to the small percentage of this group, they were treated as missing data. Educational level included 3 groups: group 1 (with high school diploma), group 2 (with associate or college degree), and group 3 (with master’s or doctoral degree). There were 3 professional groups: group 1 (practitioners, including healthcare providers and public health professionals), group 2 (researchers who use health indicator data and other researchers), and group 3 (other professional groups) (Table 1).

**Table 1.** Numbers and percentages of all independent variable groups

| Independent Variable | Group | Number | Percentage |
| --- | --- | --- | --- |
| Age | 1 (≤ 35 years) | 212 | 26.8% |
|  | 2 (36–55 years) | 242 | 30.6% |
|  | 3 (≥ 56 years) | 337 | 42.6% |
| Gender | 1 (Female) | 604 | 77.3% |
|  | 2 (Male) | 177 | 22.7% |
| Professional Group | 1 (Practitioner) | 210 | 26.5% |
|  | 2 (Researcher) | 189 | 23.9% |
|  | 3 (Other) | 392 | 49.6% |
| Education | 1 (High School) | 82 | 10.4% |
|  | 2 (College Degree) | 346 | 43.7% |
|  | 3 (Graduate Degree) | 363 | 45.9% |

Note: N = 791.

To examine the bivariate relationships of all variables, we conducted a correlation analysis among the 4 demographic factors and the 13 health indicators (Table 2). To compare the group mean differences, we performed a complete multivariate analysis of variance (MANOVA) with the 4 demographic factors as the independent variables and the 13 health indicators as the dependent variables. As a result, we identified a 2-way interaction effect between age and professional group. Therefore, the final MANOVA model included the main effects of age, gender, education, professional group, and an interaction effect between age and professional group. The null hypothesis was that a combination of the ratings of the 13 health indicators has no significant mean difference between the groups. The alpha level was set as .05 for the 4-way MANOVA analysis. The final total sample size was 791 due to the missing data from Ohio University for 5 of the 13 indicators.

**Table 2.** Correlation matrix of all variables in the study

|  | 1 | 2 | 3 | 4 | 5 | 6 | 7 | 8 | 9 | 10 | 11 | 12 | 13 | 14 | 15 | 16 | 17 |
| --- | --- | --- | --- | --- | --- | --- | --- | --- | --- | --- | --- | --- | --- | --- | --- | --- | --- |
| 1 Age | – |  |  |  |  |  |  |  |  |  |  |  |  |  |  |  |  |
| 2 Gender | .09<br>* | – |  |  |  |  |  |  |  |  |  |  |  |  |  |  |  |
| 3 Professional Group | .07<br>* | .10* | – |  |  |  |  |  |  |  |  |  |  |  |  |  |  |
| 4 Education | .12<br>* | .03 | –.16<br>* | – |  |  |  |  |  |  |  |  |  |  |  |  |  |
| 5 Immunization | .03 | –.01 | .06 | –.02 | – |  |  |  |  |  |  |  |  |  |  |  |  |
| 6 Personal Care Needs | .03 | –.04 | –.01 | –.04 | .30* | – |  |  |  |  |  |  |  |  |  |  |  |
| 7 Cancer Screening | .08<br>* | –.06 | .06 | –.11<br>* | .41* | .36* | – |  |  |  |  |  |  |  |  |  |  |
| 8 HIV Testing | –.0<br>1 | –.15<br>* | .03 | –.08<br>* | .32* | .39* | .61* | – |  |  |  |  |  |  |  |  |  |
| 9 Self-rated Health | .15<br>* | –.05 | –.00 | –.05 | .18* | .33* | .23* | .23* | – |  |  |  |  |  |  |  |  |
| 10 Air Quality Index | .14<br>* | –.04 | .04 | .02 | .29* | .40* | .36* | .36* | .31* | – |  |  |  |  |  |  |  |
| 11 Dentist Supply | .18<br>* | –.12<br>* | .05 | –.05 | .35* | .44* | .46* | .41* | .37* | .62* | – |  |  |  |  |  |  |
| 12 Health Literacy | .01 | –.09<br>* | –.03 | .03 | .28* | .39* | .31* | .33* | .29* | .49* | .48* | – |  |  |  |  |  |
| 13 Blood Sugar | .21<br>* | –.08<br>* | –.01 | –.02 | .31* | .36* | .46* | .41* | .28* | .33* | .41* | .32* | – |  |  |  |  |
| 14 Blood Triglycerides | .13<br>* | –.06 | .04 | –.05 | .35* | .36* | .52* | .44* | .26* | .32* | .40* | .32* | .73* | – |  |  |  |
| 15 HDL | .08<br>* | –.05 | .06 | –.07 | .36* | .36* | .54* | .45* | .24* | .31* | .39* | .34* | .65* | .82* | – |  |  |
| 16 LDL | .12<br>* | –.05 | .05 | –.00 | .37* | .34* | .54* | .42* | .25* | .32* | .39* | .31* | .67* | .77* | .82* | – |  |
| 17 Total Cholesterol | .07 | –.05 | .09* | –.10<br>* | .35* | .30* | .49* | .39* | .25* | .25* | .38* | .29* | .57* | .68* | .72* | .76* | – |
| *Means* |  |  |  |  | 7.52 | 6.86 | 7.25 | 6.64 | 6.67 | 6.76 | 6.47 | 7.03 | 7.74 | 7.32 | 7.32 | 7.45 | 7.25 |
| *Standard Deviations* |  |  |  |  | 2.15 | 2.08 | 2.07 | 2.36 | 2.17 | 1.92 | 2.02 | 2.02 | 1.65 | 1.80 | 1.84 | 1.86 | 2.00 |

Note: N = 791; \* indicates $p < .05$ . HDL, high-density lipoprotein; LDL, low-density lipoprotein.

This study was approved by the Institutional Review Boards of Ohio University (17-X-142) and Clemson University (IRB2019-441). All participants checked the consent form before answering the survey.

## Results

A 4-way MANOVA with *post-hoc* testing was conducted to examine mean differences among 4 independent variables (age, gender, professional group, and educational level) and a combination of dependent variables (13 health indicators). The percentages and numbers of all 4 independent variable levels are shown in Table 1 to provide a basic profile of the survey respondents. The bivariate correlations of age, gender, professional group, education level, and the 13 health indicators are shown in Table 2. The correlation results indicated that all the dependent variables were significantly correlated with each other (*Spearman’s rank correlation coefficient*, .18 –.82). The independent variable age was correlated with most of the dependent variables except for immunization, personal care needs, HIV testing, health literacy, and total cholesterol. Gender was correlated with HIV testing, dentist supply, health literacy, and blood sugar level. Professional group was correlated with total cholesterol. Education level was correlated with cancer screening, HIV testing, and total cholesterol.

The MANOVA modeling results indicated that gender, age, and education level had significant main effects on the combination of 13 health indicators, after controlling for other predictor effects. In contrast, the professional group did not have a significant main effect on the combination of 13 dependent variables after controlling for other predictor effects. However, there was a significant interaction effect of professional group with age after controlling for other predictor effects. The MANOVA results are shown in Table 3, and include Wilks’ lambda value, df, F, p-value, and partial eta squared (i.e., effect size).

**Table 3.** MANOVA results of the multivariate tests on the main effects of the 4 demographic factors on the combination of the 13 health indicators

| Effects | Wilks' Lambda value | df | F | p | Partial Eta squared |
| --- | --- | --- | --- | --- | --- |
| Gender* | 0.96 | 13 | 2.57 | <.01 | 0.043 |
| Education* | 0.94 | 26 | 1.82 | <.01 | 0.031 |
| Age* | 0.87 | 26 | 4.23 | <.001 | 0.068 |
| Professional Group | 0.95 | 26 | 1.47 | 0.06 | 0.025 |
| Age*Professional Group* | 0.90 | 52 | 1.53 | <.01 | 0.026 |

Note: N = 791; \* indicates $p < .05$ .

More specifically, the MANOVA results of the between-subjects effects (Table 4) indicated gender had a significant main effect on the respondents’ ratings of HIV testing (mean difference = .85, p < .01, partial eta squared = .023), dentist supply (mean difference = .70, p < .01, partial eta squared = .022), health literacy (mean difference = .44, p < .05, partial eta squared = .008), blood sugar level (mean difference = .41, p < .01, partial eta squared = .011), blood triglycerides (mean difference = .34, p < .05, partial eta squared = .006), HDL (mean difference = .33, p < .05, partial eta squared = .006), and LDL (mean difference = .33, p < .05, partial eta squared = .006), after controlling for other predictor effects. All the above mean differences indicated that female ratings were higher than male ratings.

**Table 4.** MANOVA results of between-subjects effects

| IVs | DVs | df | F | p | Partial Eta squared |
| --- | --- | --- | --- | --- | --- |
| Gender | Immunization/Vaccination | 1 | 0.28 | >.05 | 0.000 |
|  | Personal care needs | 1 | 1.60 | >.05 | 0.002 |
|  | Cancer screening detection | 1 | 3.66 | >.05 | 0.005 |
|  | HIV testing* | 1 | 17.57 | <.01 | 0.023 |
|  | Self-rated health status | 1 | 3.46 | >.05 | 0.005 |
|  | Air quality index > 100 | 1 | 3.19 | >.05 | 0.004 |
|  | Dentist supply* | 1 | 16.82 | <.01 | 0.022 |
|  | Health literacy rate* | 1 | 6.44 | 0.01 | 0.008 |
|  | Blood sugar level* | 1 | 8.50 | <.01 | 0.011 |
|  | Blood triglycerides* | 1 | 4.92 | 0.03 | 0.006 |
|  | HDL cholesterol* | 1 | 4.36 | 0.04 | 0.006 |
|  | LDL cholesterol* | 1 | 4.25 | 0.04 | 0.006 |
|  | Total cholesterol | 1 | 3.84 | >.05 | 0.005 |
| Education | Immunization/Vaccination | 2 | 0.33 | >.05 | 0.001 |
|  | Personal care needs | 2 | 1.19 | >.05 | 0.003 |
|  | Cancer screening detection* | 2 | 5.23 | 0.01 | 0.014 |
|  | HIV testing | 2 | 1.87 | >.05 | 0.005 |
|  | Self-rated health status | 2 | 3.14 | >.05 | 0.008 |
|  | Air quality index > 100 | 2 | 0.36 | >.05 | 0.001 |
|  | Dentist supply | 2 | 2.62 | >.05 | 0.007 |
|  | Health literacy rate | 2 | 0.86 | >.05 | 0.002 |
|  | Blood sugar level | 2 | 1.29 | >.05 | 0.003 |
|  | Blood triglycerides | 2 | 1.74 | >.05 | 0.005 |
|  | HDL cholesterol | 2 | 2.31 | >.05 | 0.006 |
|  | LDL cholesterol | 2 | 0.36 | >.05 | 0.001 |
|  | Total cholesterol* | 2 | 4.41 | 0.01 | 0.011 |
| Age | Immunization/Vaccination | 2 | 0.13 | >.05 | 0.000 |
|  | Personal care needs | 2 | 2.49 | >.05 | 0.007 |
|  | Cancer screening detection | 2 | 2.92 | >.05 | 0.008 |
|  | HIV testing | 2 | 0.70 | >.05 | 0.002 |
|  | Self-rated health status* | 2 | 10.92 | <.01 | 0.028 |
|  | Air quality index > 100* | 2 | 13.93 | <.01 | 0.035 |
|  | Dentist supply* | 2 | 17.67 | <.01 | 0.044 |
|  | Health literacy rate | 2 | 0.61 | >.05 | 0.002 |
|  | Blood sugar level* | 2 | 19.15 | <.01 | 0.048 |
|  | Blood triglycerides* | 2 | 6.67 | <.01 | 0.017 |
|  | HDL cholesterol | 2 | 2.36 | >.05 | 0.006 |
|  | LDL cholesterol* | 2 | 5.24 | <.01 | 0.014 |
|  | Total cholesterol | 2 | 1.84 | >.05 | 0.005 |
| Professional | Immunization/Vaccination | 2 | 1.47 | 0.23 | 0.004 |
|  | Personal care needs | 2 | 0.38 | 0.68 | 0.001 |
|  | Cancer screening detection | 2 | 1.37 | 0.26 | 0.004 |
|  | HIV testing | 2 | 1.34 | 0.26 | 0.004 |
|  | Self-rated health status | 2 | 1.90 | 0.15 | 0.005 |
|  | Air quality index > 100 | 2 | 0.96 | 0.38 | 0.003 |
|  | Dentist supply | 2 | 2.13 | 0.12 | 0.006 |
|  | Health literacy rate | 2 | 3.37 | 0.05 | 0.009 |
|  | Blood sugar level | 2 | 0.26 | 0.77 | 0.001 |
|  | Blood triglycerides | 2 | 0.59 | 0.56 | 0.002 |
|  | HDL cholesterol | 2 | 1.61 | 0.20 | 0.004 |
|  | LDL cholesterol | 2 | 1.26 | 0.28 | 0.003 |
|  | Total cholesterol | 2 | 2.62 | 0.07 | 0.007 |
| Age*Professional | Immunization/Vaccination | 4 | 0.67 | >.05 | 0.004 |
|  | Personal care needs* | 4 | 2.54 | 0.04 | 0.013 |
|  | Cancer screening detection | 4 | 1.13 | >.05 | 0.006 |
|  | HIV testing | 4 | 1.90 | >.05 | 0.010 |
|  | Self-rated health status | 4 | 0.33 | >.05 | 0.002 |
|  | Air quality index > 100* | 4 | 4.66 | <.01 | 0.024 |
|  | Dentist supply* | 4 | 3.04 | 0.02 | 0.016 |
|  | Health literacy rate* | 4 | 2.50 | 0.04 | 0.013 |
|  | Blood sugar level | 4 | 1.15 | >.05 | 0.006 |
|  | Blood triglycerides | 4 | 1.87 | >.05 | 0.010 |
|  | HDL cholesterol | 4 | 2.64 | >.05 | 0.014 |
|  | LDL cholesterol* | 4 | 2.74 | 0.03 | 0.014 |
|  | Total cholesterol | 4 | 1.83 | >.05 | 0.010 |

*Note:* N = 773; df of Error = 761, \* and yellow highlights indicate p < .05.

In contrast, education had a significant main effect on the respondents’ ratings of cancer screening detection (partial eta squared = .014) and total cholesterol (partial eta squared = .011), after controlling for other predictor effects. Tukey’s honest significant difference (HSD) *post hoc* results indicated that respondents with graduate degrees rated cancer screening detection approximately .65 lower than those with high school diplomas (p < .05, 95% CI = [.04, 1.25]) and approximately .44 lower than respondents with college degrees (p < .05, 95% CI = [.08, .81]). Meanwhile, respondents with graduate degrees rated total cholesterol approximately .62 lower than respondents with high school diplomas (p < .05, 95% CI = [.04, 1.21]), and approximately .41 lower than respondents with college degrees (p < .05, 95% CI = [.06, .77]).

Age had a significant main effect on the respondents’ ratings of self-rated health status, blood sugar level, and blood triglycerides and had significant simple main effects on the ratings of air quality index > 100, dentist supply, and LDL with interaction with professional group, after controlling for other predictor effects. Tukey’s HSD *post hoc* results indicated that the self-rated health status of age group 3 was approximately .76 (p < .01, 95% CI = [.31, 1.20]) higher than that of age group 1 (age ≤ 35 years) and approximately .44 (p < .05, 95% CI [.02, .87]) higher than that of age group 2 (age between 36 and 55 years). Furthermore, age group 3 rated blood sugar approximately .84 (p < .01, 95% CI = [.50, 1.17]) higher than age group 1 and approximately .38 (p < .05, 95% CI = [.06, .70]) higher than age group 2. Age group 2 rated blood sugar .46 (p < .01, 95% CI = [.10, .82]) higher than age group 1. Age group 3 rated blood triglycerides approximately .56 (p < .01, 95% CI = [.19, .93]) higher than age group 1.

The professional groups did not have a significant simple main effect on any indicator; however, the interaction effects of professional group and age were significant for the ratings of personal care needs, air quality index > 100, dentist supply, health literacy, and LDL, after controlling for other predictor effects. More details of the interaction effects are as follows.

There was a statistically significant interaction between the effects of age and professional group in personal care needs ratings (F(4,761) = 2.54, p < .05, partial eta squared = .013) (Figure 1). Bonferroni-adjusted comparisons indicated that age effects on ratings of personal care needs varied by professional group. Specifically, in professional group 1, age group 3 rated personal care needs approximately 1.15 higher than age group 1 (p < .01, 95% CI = [.24 to 2.05]), and age group 2 rated personal care needs approximately .92 higher than age group 1 (p < .05, 95% CI = [.00, 1.85]); however, in professional groups 2 and 3, the ratings did not differ with age.

**Figure 1.**
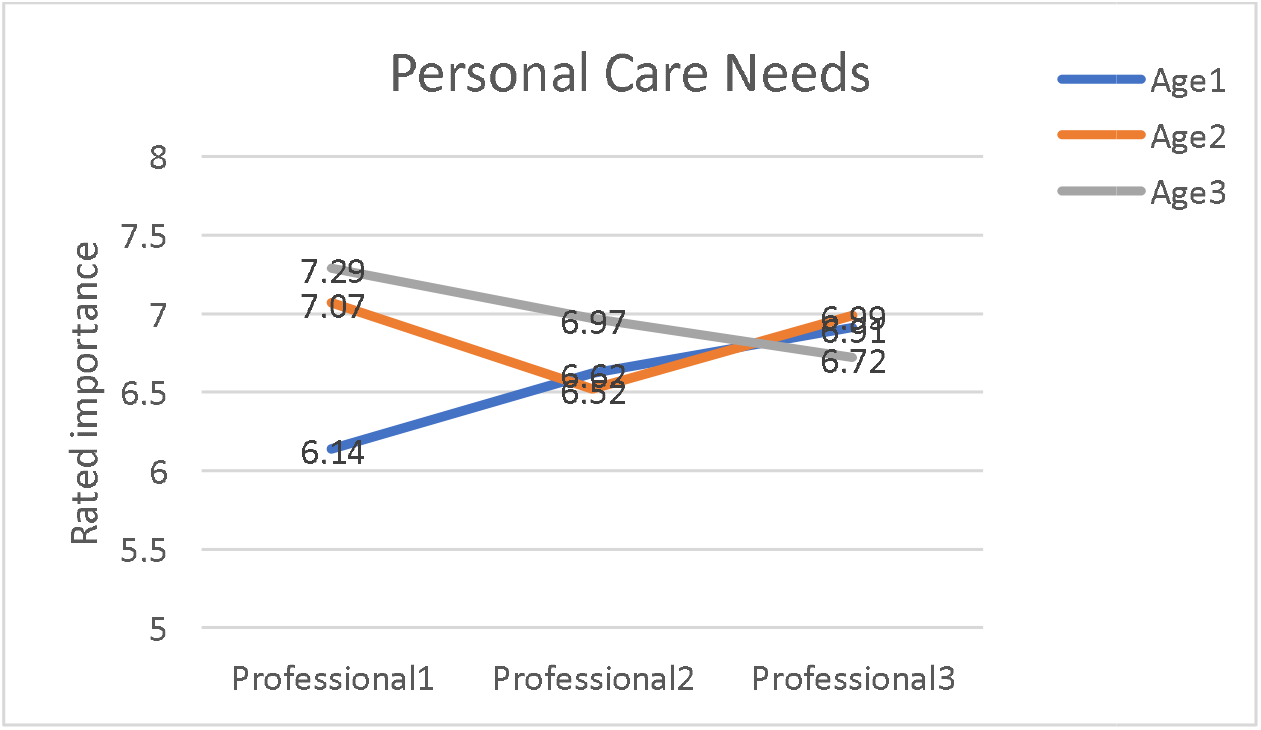
Interaction Effects of Age*Professional Group on Personal Care Needs (Age 1, ≤ 35 years; Age 2, 36– 55 years; Age 3, ≥ 56 years; Professional 1, practitioners; Professional 2, researchers; Professional 3, others)

There was a statistically significant interaction between the effects of age and professional group on air quality ratings (F (4,761) = 4.67, p < .01, partial eta squared = .024) (Figure 2). Bonferroni-adjusted comparisons indicated that age effects on ratings of air quality varied by professional group. Specifically, in professional group 2, age group 3 rated air quality approximately 1.47 higher than age group 1 (p < .001, 95% CI = [.69 to 2.24]) and approximately 1.78 higher than age group 2 (p < .001, 95% CI = [.92, 2.63]). The other two professional groups demonstrated no significant mean differences in age in terms of ratings. In contrast, the professional group ratings of air quality varied by age group. In age group 2, professional group 3 rated air quality approximately .93 higher than professional group 2 (p < .05, 95% CI = [.15, 1.70]). In age group 3, professional group 2 rated air quality approximately .90 higher than professional group 1 (p < .05, 95% CI = [.15, 1.66], and .83 higher than respondents in professional group 3 (p < .01, 95%CI = [.18, 1.48]). In age group 1, the ratings did not differ by professional group.

**Figure 2.**
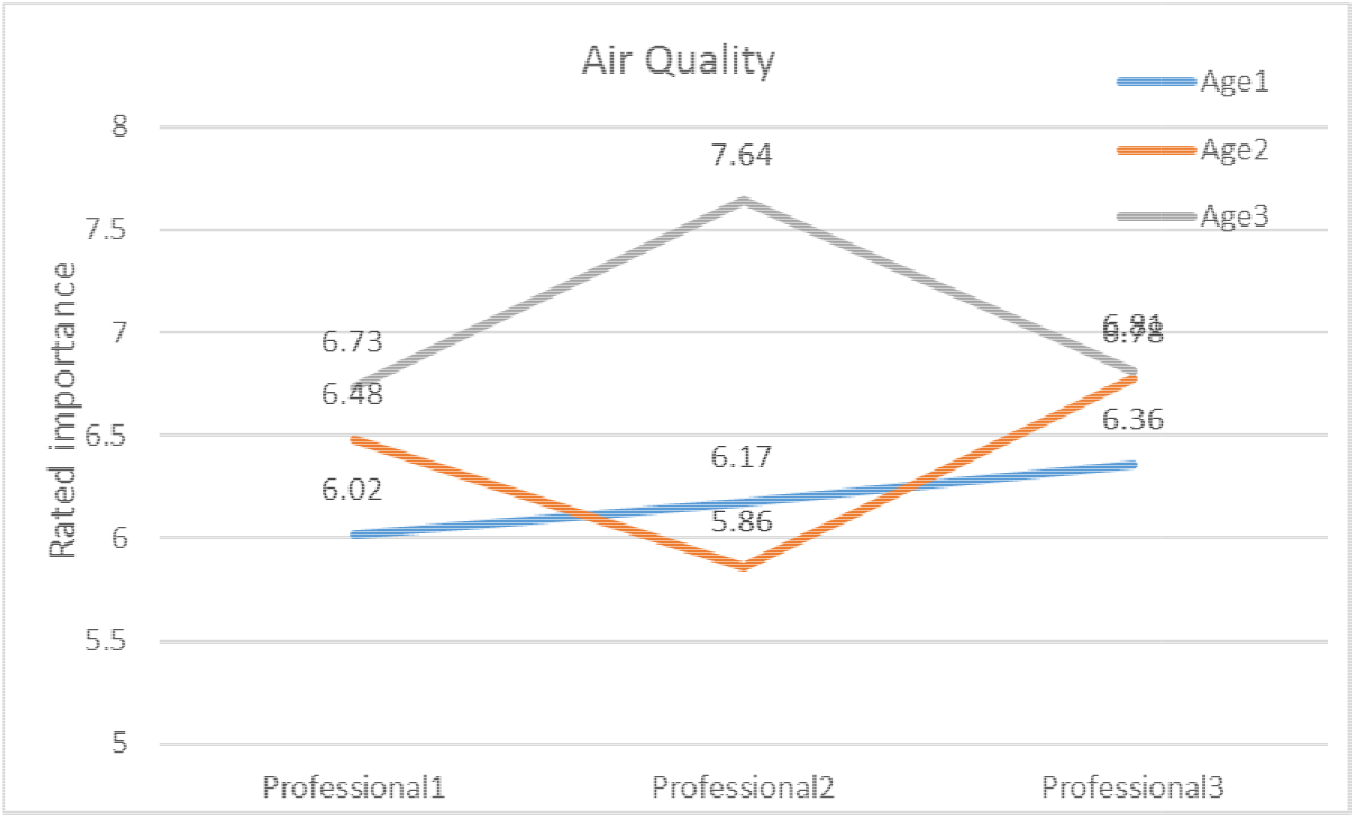
Interaction Effects of Age*Professional Group on Air Quality Index (Age 1, ≤ 35 years; Age 2, 36–55 years; Age 3, ≥ 56 years; Professional 1, practitioners; Professional 2, researchers; Professional 3, others)

There was a statistically significant interaction between the effects of age and professional group on dentist supply ratings (F(4,761) = 3.04, p < .05, partial eta squared = .016) (Figure 3). Bonferroni-adjusted comparisons indicated that age effects on the ratings of dentist supply varied by professional group. Specifically, in professional group 1, age group 3 rated dentist supply approximately 1.31 higher than age group 1 (p < .001, 95% CI = [.45 to 2.17]), and age group 2 rated dentist supply approximately 1.17 higher than age group 1 (p < .05, 95% CI = [.30, 2.04]). In professional group 2, age group 3 rated dentist supply approximately 1.14 higher than age group 1 (p < .01, 95% CI = [.34 to 1.95]) and approximately 1.41 higher than age group 2 (p < .001, 95% CI = [.53, 2.30]). In professional group 3, age group 3 rated dentist supply approximately .85 higher than age group 1 (p < .01, 95% CI = [.22 to 1.47]) and respondents in age group 2 rated dentist supply approximately .69 higher than age group 1 (p < .05, 95% CI = [.01, 1.37]). These rating differences were significantly different across the professional groups in the interaction effect. In contrast, the professional group ratings of dentist supply varied by age group. Especially in age group 2, professional group 3 rated dentist supply approximately 1.09 higher than professional group 2 (p < .01, 95% CI = [.28, 1.89]), and professional group 1 rated dentist supply approximately 1.00 higher than professional group 2 (p < .05, 95% CI = [.14, 1.86]); however, for age groups 1 and 3, the ratings did not differ by professional group.

**Figure 3.**
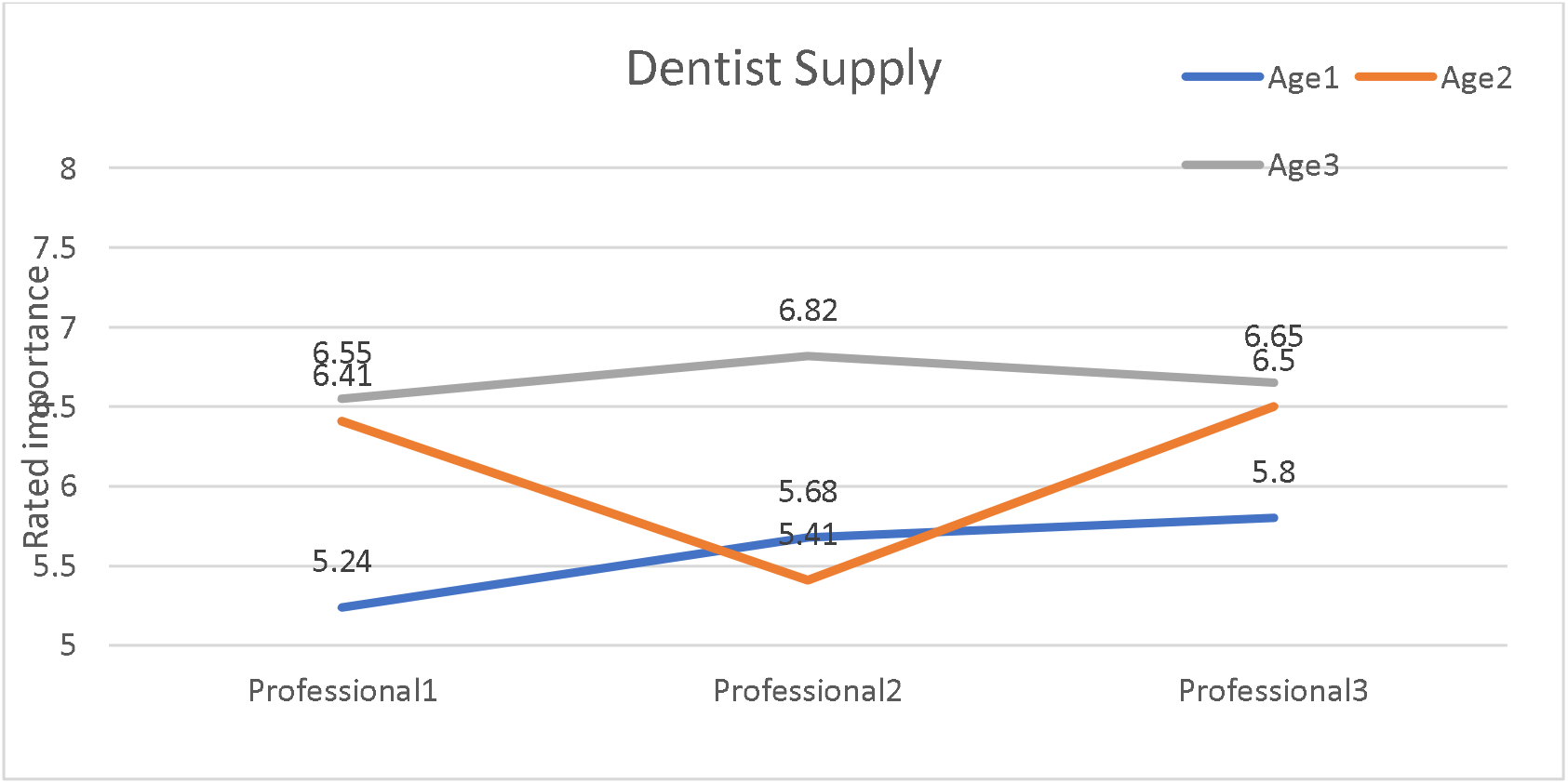
Interaction Effects of Age*Professional Group on Dentist Supply (Age 1, ≤ 35 years; Age 2, 36–55 years; Age 3, ≥ 56 years; Professional 1, practitioners; Professional 2, researchers; Professional 3, others)

There was a statistically significant interaction between the effects of age and professional group on health literacy ratings (F(4,761) = 2.50, p < .05, partial eta squared = .013) (Figure 4). Bonferroni-adjusted comparisons indicated that professional group effects on ratings of health literacy varied by age group. Specifically, in age group 2, professional group 1 rated health literacy approximately 1.28 higher than professional group 2 (p < .01, 95% CI = [.40 to 2.16]), and professional group 3 rated health literacy approximately .83 higher than professional group 2 (p < .05, 95% CI = [.01, 1.65]). In age groups 1 and 3, the ratings did not differ by professional group.

**Figure 4.**
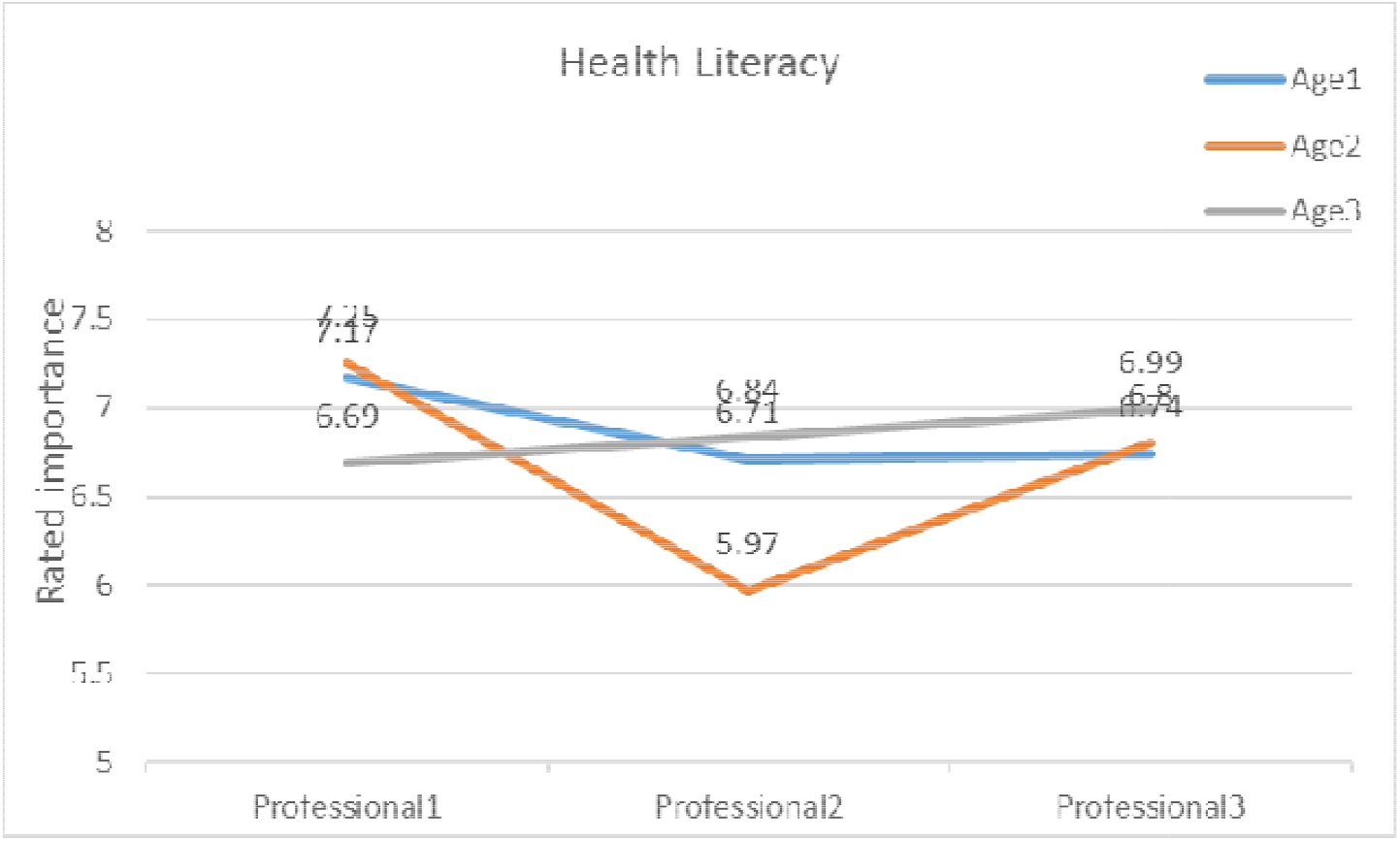
Interaction Effects of Age*Professional Group on Health Literacy (Age 1, ≤ 35 years; Age 2, 36–55 years; Age 3, ≥ 56 years; Professional 1, practitioners; Professional 2, researchers; Professional 3, others)

There was a statistically significant interaction between the effects of age and professional group on LDL ratings (F(4,761) = 2.74, p < .05, partial eta squared = .014) (Figure 5). Bonferroni-adjusted comparisons indicated that the age effects on LDL ratings varied by professional group. Specifically, in professional group 1, age group 3 rated LDL approximately .85 higher than age group 1 (p < .05, 95% CI = [.04 to 1.65]), and age group 2 rated LDL approximately .94 higher than age group 1 (p < .05, 95% CI = [.13, 1.76]). In professional group 2, age group 3 rated LDL approximately .94 higher than age group 2 (p < .05, 95% CI = [.11 to 1.76]). In professional group 3, the ratings did not differ by age group.

**Figure 5.**
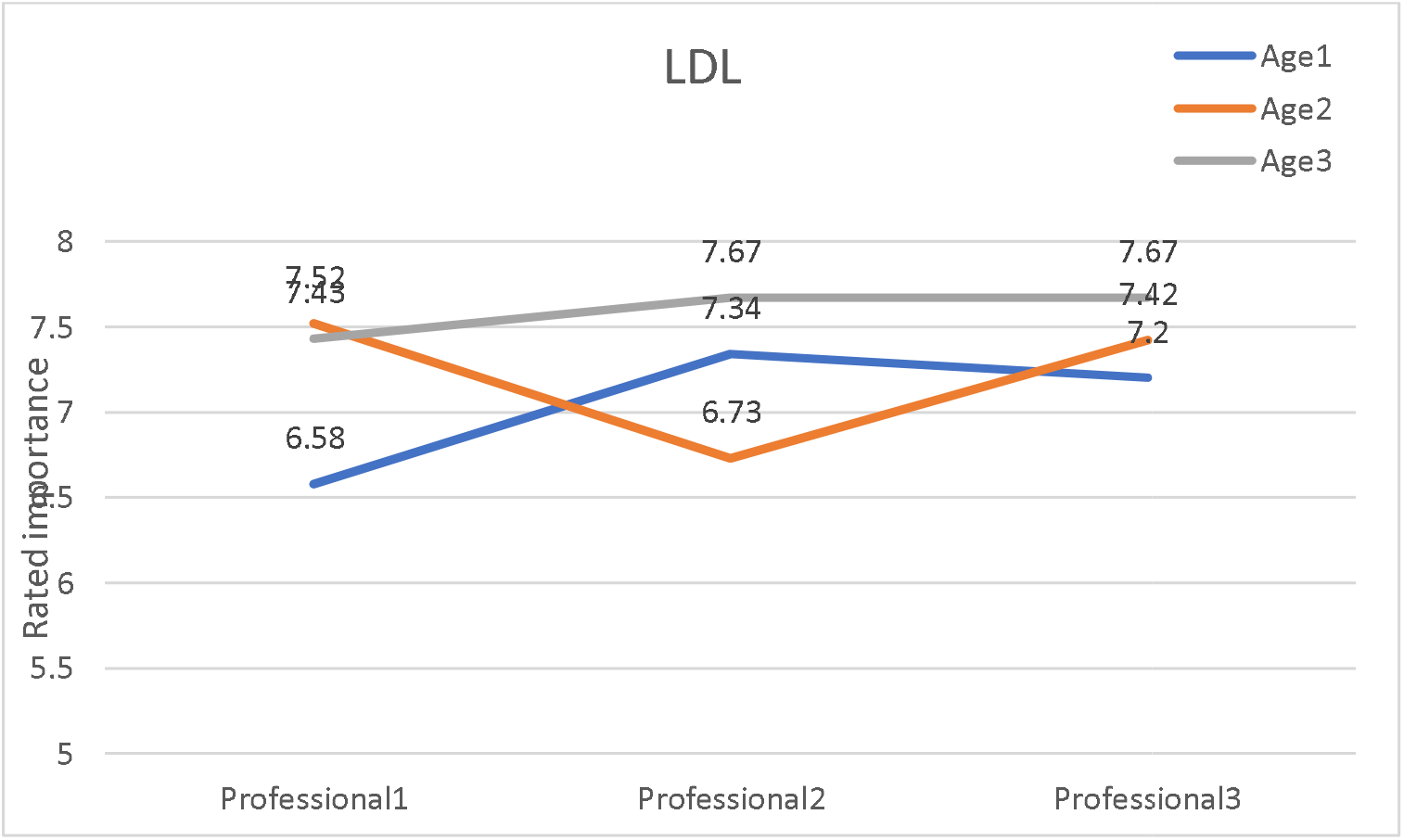
Interaction Effects of Age*Professional Group on LDL (Age 1, ≤ 35 years; Age 2, 36–55 years; Age 3, ≥ 56 years; Professional 1, practitioners; Professional 2, researchers; Professional 3, others)

## Discussion

### Interpretations of our analysis results

Our analysis indicates that the importance ratings of health indicators are not entirely independent of age, gender, professional group, or educational level. Based on the bivariate correlation results (Table 2), age correlated significantly with 8 of 13 health indicators, showing that age is a critical factor in the importance ratings. The MANOVA model (Table 3) also demonstrated that, compared with the other predictor effects, age had the largest effect on the combination of 13 health indicators, with the largest partial eta squared value (0.068) after controlling for all other variables in the model. Another age-related trend was the importance of self-related health status, blood sugar, and blood triglycerides; the older the respondents, the more important they rated these 3 health indicators.

The 4-way MANOVA model results (Table 4) indicated that gender had significant main effects on the combination of 13 health indicators when controlling for other variables. The largest gender effect was on the importance of HIV testing, implying that when considering all the variables in the MANOVA model, gender plays a critical role in determining the importance of HIV testing as a health indicator. Among all the health indicators, age had a significant main effect (Table 4), with the largest effects on dentist supply and blood sugar level ratings. Meanwhile, the educational level had a main effect on the ratings of both cancer screening detection and total cholesterol. However, the educational level had a larger effect on the cancer screening detection rating. Interestingly, the importance of cancer screening detection is inversely associated with educational level, i.e., cancer screening detection importance was rated in descending order by respondents with high school diplomas, college degrees, and graduate degrees. One possible explanation could be the perceived terminal nature of cancers and the inability of individuals to contribute to the rating patterns observed.

There were additional interesting results when examining the statistically significant interactions between age and professional group in terms of 5 health indicators (Figures 1–5). First, age group 3 rated the air quality index > 100, dentist supply, LDL, personal care needs, and health literacy indicators highest compared with the other 2 age groups across all 3 professional groups. Second, among these 5 health indicators, the interactions between age and professional group had the largest effect on the importance rating of air quality index > 100 compared with the other 4 health indicators, although all 5 were statistically significant. Third, air quality index > 100 (Figure 2) and dentist supply (Figure 3) had a similar pattern, i.e., age group 3 in professional group 2 (researchers) has the highest importance ratings and age group 2 (36 – 55) in professional group 2 (researchers) had the lowest importance ratings for the two health indicators, indicating that, among the researchers, age groups 2 and 3 showed considerable variations in the perceived importance of these health indicators. Fourth, age group 2 (36 – 55 years) of professional group 2 (researchers) had a similar rating pattern for the 5 health indicator, rating from lowest compared with other age groups or professional groups, a pattern that is observable in Figures 1 to 5 as a V-shaped orange curve.

### Significance of the work

Efforts were made to prioritize the commonly used 29 health indicators via public perspective surveys to obtain ratings on the importance of the 29 health indicators. The rating results provide a foundation to prioritize these 29 health indicators, 13 of which received relatively consistent ratings among the 3 samples. The detailed results can be used to determine which health indicators should be included in EHR systems or personal health records (PHR) systems, given that all 29 can be a burden to collect. The survey results[12] and further analysis presented in this paper provide a foundation for developing a comprehensive health indicator formula, which can measure individual health and the outcomes of preventive medicine services over time [7,16-18]. Preventive services have been recognized as important for controlling the increasing costs of healthcare [19,20]. However, the accurate measurement of such services can be challenging, given that most outcomes of such services are not measurable within a short time span.

The current paper further explored demographic factors, including age, gender, educational level, and professional group, and their effects on rating these 13 health indicators. The results showed that age is a critical factor. Gender and educational level, as well as the interactions between age and professional group, had a statistically significant main effect on these health indicators. The detailed comparison and the results based on our analysis can be used to provide evidence for future health-related study designs. The assumption is that the public’s perspective on these health indicators could influence their health-related behaviors, either consciously or subconsciously. Our study results could shed some light on the perspectives of various populations on their individual health, to design more accurate behavioral-related interventions to improve health specific to subgroups of the general population. For example, for the group of researchers aged 36–55 years, a more targeted promotion of air quality index > 100, health literacy, LDL, and dentist supply could be used to improve their health behaviors.

### Compared with other similar measures

The United Kingdom (UK) Health Index is used to track changes over time and compare the health of residents in different areas of England [2,4,21] and consists of 3 categories: healthy people, healthy lives, and healthy places. The healthy lives category is very similar to many of our health indicators, also referred to as modifiable risk factors, at the individual level. The healthy people category measures health at the population level, whereas the healthy places category measures health at the system level or beyond individual control (e.g., air quality) [22].

A closer comparison of the UK Health Index [4,21] and our 29 health indicators[12] showed that there was a large overlap between the two: 15 health indicators (Table 5). Certain mappings between our health indicators and those used in the UK Health Index were comparable, for example, BMI and unemployment were essentially the same health indicators used by both. Certain mappings were close but not the same (e.g., air quality and air pollution). More than half of the health indicators in our study were also used by the UK Health Index, which provides additional evidence of the validity of these health indicators, considering that the UK Health Index is a government project.

**Table 5.**
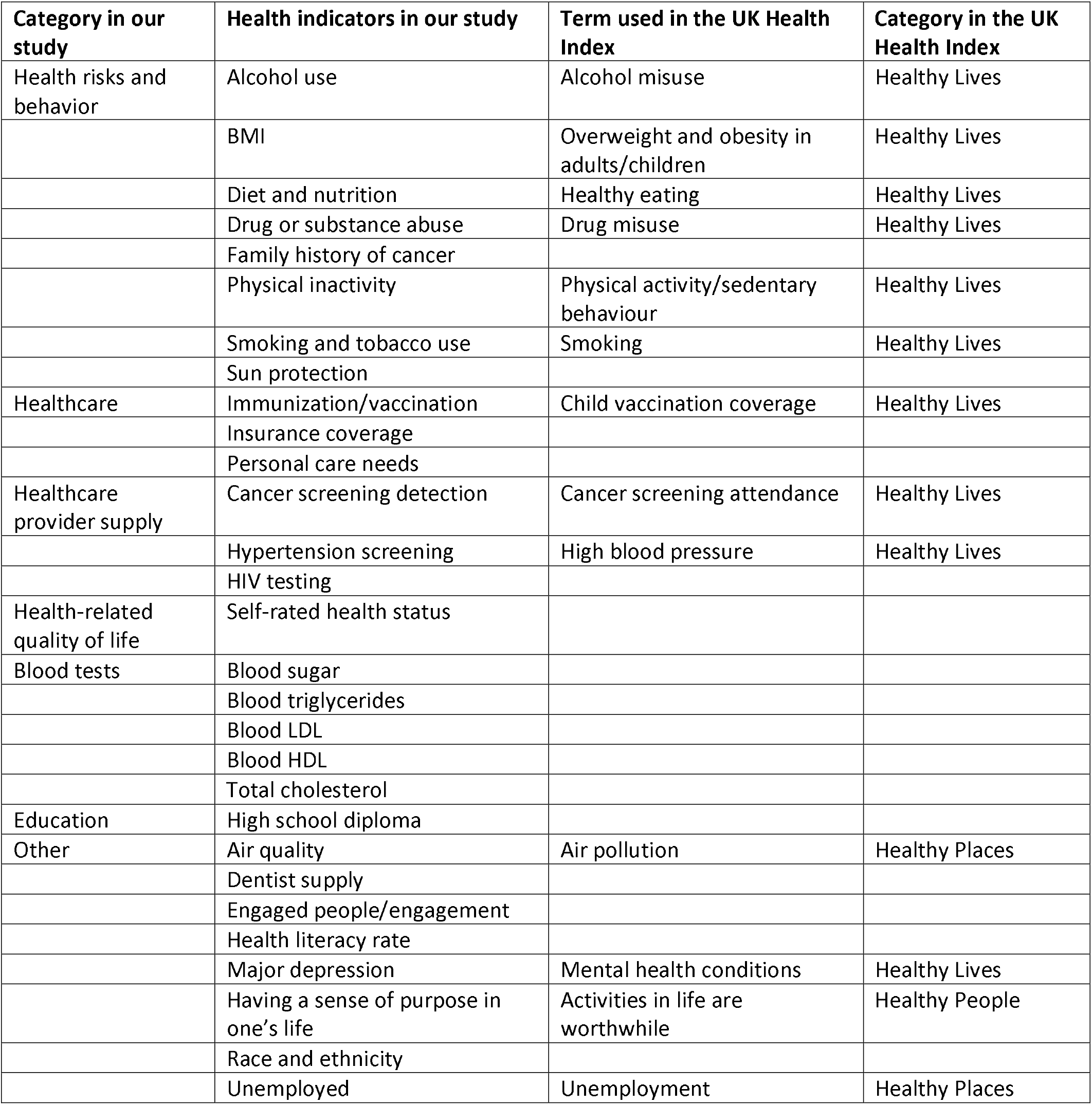
The overlap between our health indicators and the indicators used in the UK Health Index

However, after closely examining the methodology used in developing the UK Health Index, we noticed that our project differed from the UK Health Index project in the following ways: First, the UK Health Index is used to measure a nation’s health status at both the population level and individual level; our health indicators are used to measure individual health status only. Collectively, however, such indicators can be aggregated to compare different groups’ health outcomes or to track changes over time. Second, in the long-term view, our goal is to use the survey results, analysis results, and real-world data to develop a formula to calculate a more objective, data-driven health indicator to represent an individual’s overall health status. The UK Health Index project used factor analysis to group health indicators and determine the weight of each health indicator. Third, our next-stage effort will focus on formula development and the validation of the health indicators, while the UK Health Index project uses existing and available data sources and already provides Health Index comparisons among various areas in England and compares the same area over time. Although the references provided [22,23] by the UK Health Index project are very helpful, our study has a different focus and path than the UK Health Index project. Fourth, our initial goal was to measure individual health status; therefore, the validity of the measurement is our core and one long-term use scenario for our study is within EHR or PHR systems, and validity will be critical to these contexts. For the UK Health Index, however, the focus is on the availability and continuity of data sources and comparing various locations or the same location at different time points. For these purposes, the consistency of data collection across borders is much more important and is a critical contributing factor to the validity of the comparisons, given that the comparison could possibly mitigate flaws if both sides use the same criteria and methodology during data collection and analysis. However, these two projects are not in parallel, because the UK Health Index project is a government project based on rich longitudinal data sources. In contrast, ours was an investigator-initiated exploratory project. Therefore, the platforms and available resources between the projects are not comparable, despite sharing certain similarities in methodology and vision.

### Strengths and weaknesses of the study

Our study is a new exploration of how to measure individual health more objectively, accurately, and comprehensively. Currently, an objective and accurate measure of individual health status is lacking. Such a measure can track numerous operationally related healthcare system performances, preventive services for individuals, and the health and outcomes of the population over time. In addition, such measures can be used to establish more comparable groups for conducting epidemiological or health service studies. Despite our results being the first step in the development of a larger project, its direction is critical to improving the health of the whole population and the use and promotion of preventive care services. Although we have taken the initial steps in this direction, our work is still in the preliminary stage.

The main limitation of our study is that we are at the beginning of the project. Our current analysis is based on public perspectives without additional validation. Despite this reality, we feel that our deep analysis of the demographic variables of individual respondents and their effects on and interactions with the rating results are nevertheless helpful for thoroughly understanding the perspectives. In addition, this analysis and the results can be used to set the foundation for future formula development to create a comprehensive individual Health Index.

Another limitation is that our results are derived from survey results; despite the relatively large sample sizes, our results are more in line with an observational study. These results can shed light on the correlations between various demographic subgroups and ratings. However, given that they are not as convincing as those from a hypothesis-driven randomized controlled study, we urge readers to use the results within the context.

### Potential future developments

Our next step is to explore longitudinal individual health data, possibly through EHR, to establish comparable groups to further refine and validate the public survey results. The ultimate goal is to construct and validate a formula to calculate a Health Index for individuals by using weighted health indicators. Such a validated Health Index can be used to track changes, to measure operational level healthcare services more accurately and consistently, and to form more comparable control groups during epidemiological studies.

## Conclusion

All 13 health indicators were significantly intercorrelated. Our results showed that age was critical in rating the 13 health indicators. The MANOVA modeling results indicated that age, gender, and education levels had significant main effects on the combination of 13 health indicators. Age had the largest effect size, and gender had a greater effect size than educational levels, according to our MANOVA model. There was a significant interaction effect of professional group with age; however, the effect size was smaller than age, gender, and educational levels. The correlation results and the MANOVA model between demographic factors and rating results can provide a foundation for the following: 1) developing a formula for an overall Health Index; 2) providing evidence for study designs on precise behavioral interventions for individual subgroups of the general public; and 3) providing evidence for thorough understanding of the public perspectives of the indicators.

## Data Availability

The data sets and statistical analysis strategies and codes for this paper are available upon request from the corresponding author.

## Acknowledgements

None

## Author contributions

XJ and YZ determined the initial analytic strategies and YZ conducted the data analysis and drafted the Methods and Results sections. XJ and YZ drafted the initial manuscript together. TS cleaned the data and organized them in a usable manner with guidance from YZ and XJ. XJ, TS, FL, and SD were responsible for the initial conceptualization and design of the project. XJ, TS, YZ, SD, TL, LH, LS, SG, and RWG agreed on the data analysis and results interpretation and substantially contributed to revising the manuscript.

## Competing Interest

None declared.

## Funding statement

None

## Notes

### Competing Interest Statement

The authors have declared no competing interest.

## References

1. Healthy People 2020 H. Healthy People 2020 Leading Health Indicators: Progress Update, 2020.

2. LCP. UK Office for National Statistics (ONS) Health Index Explorer. Secondary UK Office for National Statistics (ONS) Health Index Explorer 2018. https://healthindex.lcp.uk.com/.

3. Medicine Io. Vital Signs: Core Metrics for Health and Health Care Progress. Washington, DC: The National Academies Press, 2015.

4. UK OfNS. UK Census 2021-Health and social care. Secondary UK Census 2021-Health and social care 2022. https://www.ons.gov.uk/peoplepopulationandcommunity/healthandsocialcare.

5. Idler EL, Benyamini Y. Self-rated health and mortality: A review of twenty-seven community studies. Journal of Health and Social Behavior 1997;38(1):21–37.

6. Mossey JM, Shapiro E. Self-rated health: A predictor of mortality among the elderly. American Journal of Public Health 1982;72(8):800–08.

7. Wold C. Health Indicators: a review of reports currently in use (2008). Secondary Health Indicators: a review of reports currently in use (2008) 2009. http://www.cherylwold.com/images/Wold_Indicators_July08.pdf.

8. National Center for Health Statistics C. National Health Interview Survey. Secondary National Health Interview Survey 2022. https://www.cdc.gov/nchs/nhis/.

9. Medicine Io. State of the USA Health Indicators: Letter Report. Washington, DC: The National Academies Press, 2009.

10. Index G-HW-B. State of global well-being: results of the Gallup-Healthways global well-being index (2014), 2015.

11. National Academies of Sciences E, Medicine. Leading Health Indicators 2030: Advancing Health, Equity, and Well-Being. Washington, DC: The National Academies Press, 2020.

12. Sokoya T, Zhou Y, Diaz S, et al. Health Indicators as Measures of Individual Health Status and Their Public Perspectives: Cross-sectional Survey Study. J Med Internet Res 2022;24(6):e38099. doi: 10.2196/38099

13. Jing X, Lekey F, Kacpura A, Jefford K. Health indicators within EHR systems in primary care settings: availability and presentation. Shanghai, China: Springer, 2016.

14. McPeek-Hinz E, Boazak M, Sexton JB, et al. Clinician Burnout Associated With Sex, Clinician Type, Work Culture, and Use of Electronic Health Records. JAMA Netw Open 2021;4(4):e215686. doi: 10.1001/jamanetworkopen.2021.5686

15. Vanderbilt University Medical Center CTSA. Research Match. Secondary Research Match 2021. https://www.researchmatch.org/about/.

16. Hensrud DD. Clinical preventive medicine in primary care: background and practice: 2. Delivering primary preventive services. Mayo Clin Proc 2000;75(3):255–64. doi: 10.4065/75.3.255

17. Hensrud DD. Clinical preventive medicine in primary care: background and practice: 3. Delivering preventive screening services. Mayo Clin Proc 2000;75(4):381–5. doi: 10.4065/75.4.381

18. Hensrud DD. Clinical preventive medicine in primary care: background and practice: 1. Rationale and current preventive practices. Mayo Clin Proc 2000;75(2):165–72. doi: 10.4065/75.2.165

19. Ozieh MN, Bishu KG, Dismuke CE, Egede LE. Trends in healthcare expenditure in United States adults with chronic kidney disease: 2002-2011. BMC Health Serv Res 2017;17(1):368. doi: 10.1186/s12913-017-2303-3

20. Cannon A, Handelsman Y, Heile M, Shannon M. Burden of Illness in Type 2 Diabetes Mellitus. J Manag Care Spec Pharm 2018;24(9-a Suppl):S5–s13. doi: 10.18553/jmcp.2018.24.9-a.s5

21. Ceely G. Health Index indicators and definitions. Secondary Health Index indicators and definitions 2022. https://www.ons.gov.uk/peoplepopulationandcommunity/healthandsocialcare/healthandwellbeing/methodologies/healthindexindicatorsanddefinitions.

22. Ceely G. Health Index methods and development: 2015 to 2019. Secondary Health Index methods and development: 2015 to 2019 2022. https://www.ons.gov.uk/peoplepopulationandcommunity/healthandsocialcare/healthandwellbeing/methodologies/healthindexmethodsanddevelopment2015to2019.

23. OECD (Organisation for Economic Co-operation and Development), The Applied Statistics and Econometrics Unit of the Joint Research Centre (JRC), The European Commission. Handbook on Constructing Composite Indicators: METHODOLOGY AND USER GUIDE, 2008.

